# Using GenAI to train mental health professionals in suicide risk assessment: Preliminary findings

**DOI:** 10.1101/2024.07.17.24310579

**Authors:** Zohar Elyoseph, Inbar Levkovitch, Yuval Haber, Yossi Levi-Belz

**Author notes:** **Corresponding Author:** Dr. Zohar Elyoseph Address: Hattena 14b, Kiryat Tivon, Israel. •. Declaration of Relevant Financial Interests The authors declare the following potential. conflicts of interest: ’None’.

## Abstract

**Background:** Suicide risk assessment is a critical skill for mental health professionals (MHPs), yet traditional training in this area is often limited. This study examined the potential of generative artificial intelligence (GenAI)-based simulator to enhance self-efficacy in suicide risk assessment among MHPs.

**Method:** A quasi-experimental study was conducted with 43 MHPs from Israel. Participants attended an online seminar and interacted with a GenAI-powered suicide risk assessment simulator. They completed pre- and post-intervention questionnaires measuring suicide risk assessment self-efficacy and willingness to treat suicidal patients. Qualitative data on user experience were collected.

**Results:** We found a significant increase in self-efficacy scores following the intervention. Willingness to treat patients presenting suicide risk increased slightly but did not reach significance. Qualitative feedback indicated that participants found the simulator engaging and valuable for professional development. However, participants raised concerns about over-reliance on AI and the need for human supervision during training.

**Conclusion:** This preliminary study suggests that GenAI-based simulators hold promise as a tool to enhance MHPs’ competence in suicide risk assessment. However, further research with larger samples and control groups is needed to confirm these findings and address ethical considerations surrounding AI use in suicide risk assessment training. AI-powered simulation tools have the potential to democratize access to high-quality training in mental health, potentially contributing to global suicide prevention efforts. However, their implementation should be carefully considered to ensure they complement rather than replace human expertise.

## Introduction

Suicide prevention is one of the most pressing and complex issues in the field of mental health. Its critical importance is emphasized as suicide poses a serious public health concern, with annual global figures indicating approximately 817,000 individuals dying by suicide and 20 million attempted suicides.^1^ These staggering figures underscore the urgent necessity for comprehensive investment in a broad array of suicide prevention strategies aimed at curtailing local and global suicide prevalence. Thus, training professionals and gatekeepers to evaluate, react, and perform early interventions becomes crucial, among other important targets. Critical actions in this context include suicide risk assessment, which is recognized as a critical step toward suicide prevention.^2^ This study aimed to examine the ability of an artificial intelligence (AI)-based simulator to enhance competence in suicide risk assessment among mental health professionals (MHPs).

Suicide risk assessment is a process in which MHPs seek to assess the likelihood of a patient engaging in future suicidal behavior.^3^ Currently, this assessment primarily includes administering the Columbia-Suicide Severity Rating Scale (C-SSRS^4^), which poses direct questions about the frequency, intensity, and content of suicidal thoughts, detailed questions about plans for future suicidal acts or descriptions of previous suicide attempts, and specific questions about the desire to die or continue living.^4^ Suicide risk assessment also includes the assessment of risk and protective factors relating to depressive symptoms and recent mental crises.^5^Despite the recent positive global trend of making knowledge about suicide risk assessment more accessible in the community, such programs remain limited. There is a dearth of experts to deliver appropriate training, and existing programs are often taught at a theoretical level without allowing intensive training with patients and frequent professional feedback. At best, MHPs usually watch a video demonstrating how to conduct an assessment but do not perform multiple assessments with diverse types of patients with various risk factors, communication styles, psychopathologies, and other difficulties. These shortcomings reflect a significant training gap for one of the most challenging clinical tasks in mental health, a task that is already considered very difficult to predict,^6^ partly due to the impulsive nature of suicidal behavior and the large variability among patients.^7^

Importantly, in several studies, MHPs were found to be highly reluctant to treat suicidal individuals,^8^ partly due to their perception of low knowledge and competence regarding suicide risk assessment.^9^ Improving the knowledge and competence of MPHs in performing real-time suicide risk assessments would be a crucial step toward accurately identifying those at risk and providing the appropriate psychological help. Thus, improving these skills would require systematic training in controlled conditions for real-time risk assessment. However, how competence can be enhanced among MHPs remains an open question. Following this, in this study, we aimed to shed light on the prospect of employing an AI-based simulator to enhance MHPs’ competence in suicide risk assessment. More generally, the current research will focus on how generative artificial intelligence (GenAI) technology can assist in global efforts to prevent suicide.

### AI-based technology in mental health

GenAI––large language models (LLMs)––is an advanced technology that entered widespread public use by late 2022. Within two months, 100 million users adopted applications such as ChatGPT,^10^ considered the fastest technology adoption rate in history. Since then, numerous studies have demonstrated the significant potential of this technology in various domains, including education, medicine, law, art, programming, and psychology.^11–14^

LLMs are a cornerstone technology within the broader GenAI field, designed to understand, generate, and manipulate human-like texts based on vast amounts of training data. While GenAI encompasses various AI systems capable of creating new content (including images, audio, and code), LLMs specifically focus on language tasks, serving as the foundation for advanced natural language processing applications and driving significant improvements in AI’s ability to interact with humans in more natural and contextually appropriate ways.

Recent research has shown that LLMs can accurately identify emotions and mental disorders, such as schizophrenia, depression, and anxiety, and provide treatment recommendations and prognoses comparable to mental health professionals.^15–27^ Despite their potential to democratize clinical knowledge and encourage ideological pluralism,^21,28,29^ ethical concerns persist. These include data privacy, algorithmic opacity, threats to patient autonomy, risks of anthropomorphism, technology access disparities, corporate concentration, deep fakes, fake news, reduced reliance on professionals, and amplification of biases.^12,21,28,29^ At a more philosophical level, questions arise regarding GenAI-human relationships, the prospect of generating intimacy, and how this may affect human-to-human relationships.^12,18,30^ One promising domain in which GenAI has the potential to make a significant contribution to mental health is a global endeavor to prevent suicide. ^22,24,31–35^

### GenAI and suicide prevention

Recent research has demonstrated GenAI’s potential in suicide prevention and mental health support. Studies have shown that GenAI, such as ChatGPT-4, can assess suicide risk with accuracy comparable to mental health professionals^24,31^ and adapt its assessments to different cultural contexts.^35^ GenAI has also proved proficient in using the World Health Organization’s guidelines to evaluate media reports on suicide.^22^ Beyond risk assessment, recent studies have highlighted the significant potential of GenAI in facilitating role-playing scenarios for educational and therapeutic purposes.^36,37^ This AI application offers a versatile and interactive platform for training and intervention. In therapeutic settings, GenAI can assist children and adolescents in sharing their experiences, organizing them coherently, identifying strengths and resilience resources, and creating visual representations of their internal psyche.^38^ Moreover, GenAI can also be utilized to externalize inner voices, allowing patients to engage in guided dialogue with different aspects of their internal psyche, all within a controlled therapeutic environment.^39^

Combining GenAI capabilities in suicide risk assessment with its application in role-playing scenarios offers new possibilities for mental health care and professional training. This integration may enable immediate and wide-scale access to clinical knowledge about suicide, be adaptable to various users and situations, and create interactive learning experiences. The current study examined this potential through a GenAI-based simulator designed to train MHPs to assess and mitigate suicide risk by utilizing role-play scenarios.

### The Current Study

This study aimed to make significant theoretical and practical contributions to the field of suicide prevention by harnessing the capabilities of GenAI to develop an innovative simulation tool for training MHPs. By addressing critical gaps in current training programs, such as the lack of opportunities for intensive practice and personalized feedback, this study aimed to democratize access to essential clinical knowledge and skills, ultimately contributing to global suicide prevention efforts. Specifically, We have developed an AI-powered simulator for suicide risk assessment training. This tool enables mental health professionals (MHPs) to practice and improve their skills by interacting with AI-generated patient characters. The simulator plays the role of an individual coming in for assessment, allowing MHPs to conduct mock interviews, evaluate suicide risk levels, and receive feedback on their performance. We examined possible changes in the MHPs’ perceived competence to conduct suicide risk assessments before and after training in the simulator.

We posited two primary research questions:

1. To what extent can the use of an AI-based suicide risk assessment simulator improve the self-efficacy and willingness of MHPs to handle suicide-related situations?
2. How can the design and feedback delivery within an AI-powered simulation tool be optimized to enhance learning outcomes and user experience?

## Method

### Participants and Study Design

#### Participants

The study included 43 mental health professionals (e.g., psychologists, social workers, and psychiatrists) at different professional and seniority levels in Israel. Participants were recruited through a social media. These professionals were invited to attend a special webinar on 6 June 2024 (via the Zoom application platform), during which the simulator was presented, followed by an active online experience with the simulator. This experience included completing a questionnaire tapping the main issues of this study at two times: once before engaging with the simulator (pre-measure) and once after engaging with the simulator (post-measure).

#### Study Design

This study integrated quantitative and qualitative approaches to evaluate the effectiveness and ethical considerations of using an AI-powered chatbot for suicide risk assessment training. As seen in Figure 1, the experimental design included pre- and post-intervention measurements.

**Figure 1:**
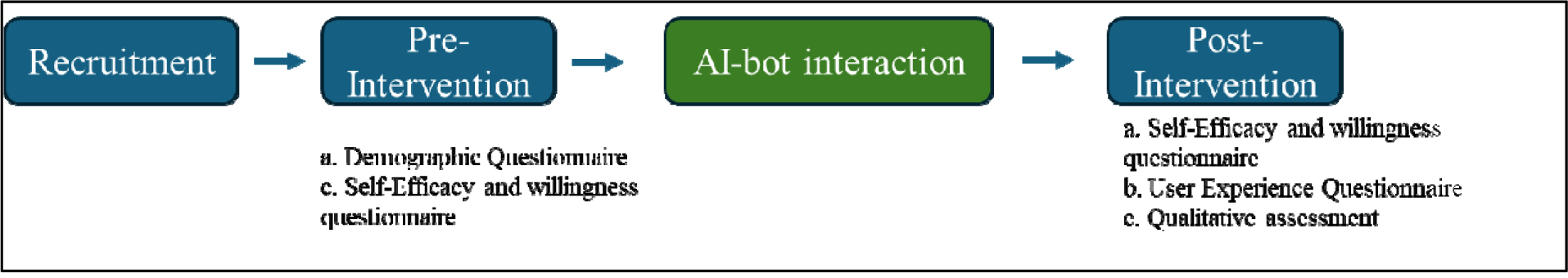
The study flow from pre- to post-intervention

#### Intervention

The Generative AI Suicide Risk Assessment bot is an innovative tool designed to enhance the training of mental health professionals in assessing suicide risk. The AI-Bot was built by the authors between March and June 2024. Interaction with the bot comprises three key stages. First, the user was introduced to the objective and structure of the simulation; the introduction provided a clear framework for the interactive learning experience. Second, the user was invited to engage in conversation (in chat interface) with one out of two unique characters, each of whom presented a distinct scenario (man or woman, a unique case story, communication style, and risk factor profile). Finally, the bot presented detailed, constructive writen feedback on the central aspects of the risk assessment procedure. This feedback helps users identify areas to improve and refine their skills.

The feedback provided in this simulation covers several critical aspects of suicide risk assessment. It evaluates the interviewer’s ability to establish rapport and trust, explores risk factors across various domains (demographic, psychological, interpersonal, and situational), identifies protective factors, and assesses the level of distress and psychopathology. The feedback also evaluates the thoroughness of suicidal thought exploration, including the nature, frequency, intensity, and duration of such thoughts, as well as any specific plans or preparatory behaviors for suicide. Additionally, the feedback offers an assessment of the interviewer’s skill in reflecting on the risk level while engaging the patient in hope and reasons for living. This comprehensive approach aligns with established suicide risk assessment protocols, such as the Columbia-Suicide Severity Rating Scale (C-SSRS)^40^. The C-SSRS provides a structured method for evaluating and improving clinical interviewing skills in high-stake mental health scenarios.

### Measures

#### Pre-Intervention Questionnaire

Participants completed a brief questionnaire assessing age, gender, profession, years of experience, and prior experience with suicide risk assessment. Participants completed a pre-intervention questionnaire designed to assess their baseline perceptions and attitudes toward conducting suicide risk assessments. The questionnaire included the following four items presented on an 11-point Likert-type scale, ranging from 0 (*not at all*) to 10 (*completely*):

Self-efficacy:

1. To what extent do you feel capable of conducting a suicide risk assessment?
2. To what extent do you feel you have the tools to conduct a suicide risk assessment?

These questions were averaged to create a composite variable called “ suicide risk assessment self-efficacy” This self-efficacy measure yielded good internal reliability (Cronbach’s alpha = .97), indicating that the items consistently measured the same underlying construct of the participants’ belief in their ability to conduct suicide risk assessments.

Willingness to treat:

1. 3. If given the opportunity, how willing would you be to take patients with suicidal risk for assessment?
2. 4. If a patient dealing with active suicidality were referred to you, how willing would you be to accept them?

These two questions were averaged to create a composite variable called “Willingness to Treat.” This measure yielded good internal reliability (Cronbach’s alpha = 0.89), indicating that the items consistently measured the same underlying construct of the participants’ willingness to treat.

#### Post-simulation questionnaire

A post-intervention questionnaire was designed to assess the participants’ experiences and attitudes following the intervention. This questionnaire included 4 items identical to those in the pre-intervention questionnaire, allowing for a direct comparison of suicide risk assessment self-efficacy (Cronbach’s alpha = .86) and willingness to conduct suicide risk assessments (Cronbach’s alpha = .90). The post-intervention questionnaire included four new items to assess the participants’ perceptions of the simulator experience:

1. To what extent did the simulator experience help you conduct future suicide risk assessments?
2. To what extent do you feel you have learned from your experience?
3. To what extent do you feel that the feedback you received will help you in future assessments?
4. Would you recommend using this AI-based simulator with other practitioners before they conduct suicide risk assessments?

Moreover, the questionnaire featured four open qualitative questions aimed at gathering deeper insights: How was your learning experience with an AI-based simulator? What do you suggest improving in the AI simulator? What advantages do you find in this training? What risks or concerns do you have about training this way?

### Procedure

The study was conducted during a live webinar (online conference) initiated by the authors, in which the participants were invited to participate in the experiment voluntarily. All questionnaires, including both pre- and post-intervention measures, as well as the demographic questions and open qualitative items, were administered through Google Forms. Participants were recruited from an MHP professional community called “The Artificial Third - Artificial Intelligence in Mental Health” and from other online communities. During the webinar, an explanation of the bot was provided. Interested participants received links via Zoom chat to pre and post intervention questionnaires and the AI-based simulator. Subsequently, they were asked to choose to interact with one of the two selected bots (male or female). Following this interaction, participants were given a post-intervention questionnaire. The interaction with the AI-based simulator took approximately 15-25 minutes. Each participant communicated with the bot from their home. Participants were instructed to engage in the interaction using a computer rather than a mobile device.

This study was approved by the University Ethics Committee (Institutional Review Board Approval Number: 2024-67 YVC EMEK). Participants were fully informed of the study’s aims, procedures, and the right to withdraw at any stage without any repercussions. Participants were assured of the confidentiality and anonymity of their data. They were instructed not to enter any personal information during interactions with the bot.

All conversations were recorded for research purposes with the participants’ informed consent. A full demonstration of one conversation is presented in Appendix A. The data were securely stored and managed by the current research team to ensure the participants’ privacy and confidentiality. The documentation of these interactions provides valuable insights into the assessment process. It enables an in-depth analysis of the MHPs’ interviewing techniques, risk evaluation strategies, and the overall effectiveness of AI-powered simulation for training MHPs.

The simulator is powered by GPT-4o (OpenAI LTD), an advanced LLM that demonstrates sophisticated and multimodal capabilities in “understanding” and generating text, recognizing, and producing speech tones, and offering improved speed for enhanced user experience. Access to this simulation system is facilitated through API technology, enabling seamless integration with external platforms. The AI bot was deployed using an innovative application––PMFM AI––which specializes in making AI-powered chatbots accessible to the public via API technology.

### Data analysis

Descriptive statistics were used to describe the participants’ demographic baseline data, followed by a series of paired *t*-tests to compare the MHPs’ responses before and after the intervention on the study measures. Pearson correlations were calculated to examine the relationships between the background data and the dependent variables. The criterion for determining statistical significance throughout the study was set at *p* < .05. Qualitative data were analyzed using content analysis.

## Results

### Information on the study sample

Participants included 43 mental health professionals with diverse backgrounds and experience levels. The participants’ ages ranged from 26 to 72 (*M*_age_ = 45.44, *SD* = 10.43). The sample’s gender distribution was mostly female, with 35 women and eight men. Regarding professional backgrounds, most participants (*n* = 27, 62.8%) were psychologists at various stages of their careers, ranging from those pursuing graduate studies to experienced supervisors. The sample also included nine social workers, one psychiatrist, and several other mental health professionals, such as a clinical criminologist, a mental health nurse, and emotional therapists. The professional status of the participants varied: 18 were experts in their field, 12 were supervisors, nine were interns, two were practicum students, and two had relevant experience (e.g., volunteering in a crisis intervention center). The participants had a wide range of professional experience in mental health, averaging 13.81 years (*SD* = 10.06), with some having as little as two years of experience and others having up to 45 years in the field. Most participants (*n* = 41, 95.34%) reported having conducted suicide risk assessments in the past. All participants indicated that they had worked with patients exhibiting suicidal characteristics, including suicidal ideation, self-injury, suicidal behavior, or past suicide attempts. Regarding prior training in suicide prevention, most participants (*n* = 29, 67.44%) had undergone formal training in suicide prevention (more than 10 hours). Seven participants reported having attended a lecture on the subject, while three had not received any specific training. Some participants mentioned receiving training through academic courses, military service, or as part of their professional development.

### Impact of the intervention on suicide risk assessment self-efficacy

A paired-sample *t*-test was conducted to evaluate the impact of the intervention on participants’ self-efficacy in conducting suicide risk assessments. As shown in Figure 2, self-efficacy scores on the 11-point scale increased significantly from pre-intervention (*M* = 6.0, *SD* = 2.4) to post-intervention (*M* = 6.4, *SD* = 2.1), *t*(42) = - 1.96, *p* = .027 (one-tailed). This finding indicates that the participants believed they were more capable of conducting suicide risk assessments following the intervention.

**Figure 2:**
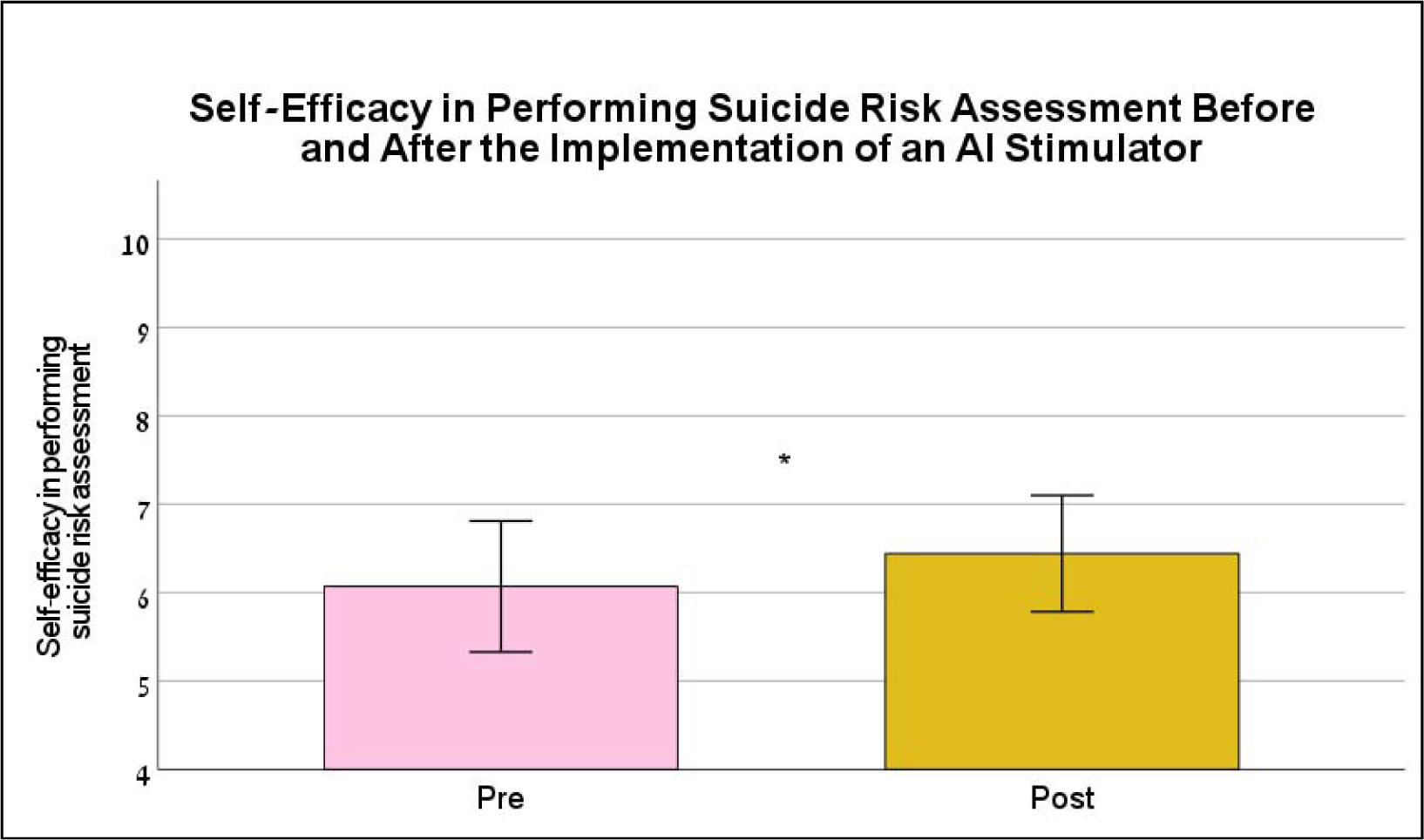
Self-efficacy in performing suicide risk assessment pre- and post-AI simulator intervention (*N* = 43) *Note.* This graph illustrates the self-efficacy of the participants in performing suicide risk assessments before and after using the AI simulator. The pink bar represents the mean pre-intervention self-efficacy score (Pre), whereas the yellow bar represents the mean post-intervention self-efficacy score (Post).*.05

### Impact of the intervention on willingness to treat

A paired-sample *t-*test was conducted to evaluate the impact of the intervention on participants’ willingness to treat patients at risk for suicide. Willingness-to-treat scores increased slightly from pre-intervention (4.76±2.64) to post-intervention (5.00±2.50), but the difference did not achieve significance, *t*(42) = -1.148, *p* = .257. The mean increase in willingness-to-treat scores was 0.24, with a 95% confidence interval ranging from .67 to .19.

### Correlation Analysis

Pre-intervention suicide risk assessment self-efficacy and willingness to treat were positively correlated (*r* = .71, *p* < .001). MHPs’ experience (in years) was positively correlated with pre-intervention self-efficacy (*r* = 0.331, *p* =.03) but not with pre-intervention willingness to treat.

### Simulator Usage Experience

As illustrated in Figure 3, most participants reported positive learning outcomes from using the simulator. For the question regarding to what extent the simulator helped in conducting future assessments, the mean score was 6.19 ± 2.26. When asked how much they felt they learned from the experience, the mean score was 6.40 ± 2.11. Regarding how helpful the feedback was for future assessments, participants reported a mean score of 6.74 ± 1.94. Lastly, for the recommendation to other practitioners, the mean score was 7.63 ± 2.27. These results suggest that participants generally found the simulator beneficial and were likely to recommend its use to others.

**Figure 3:**
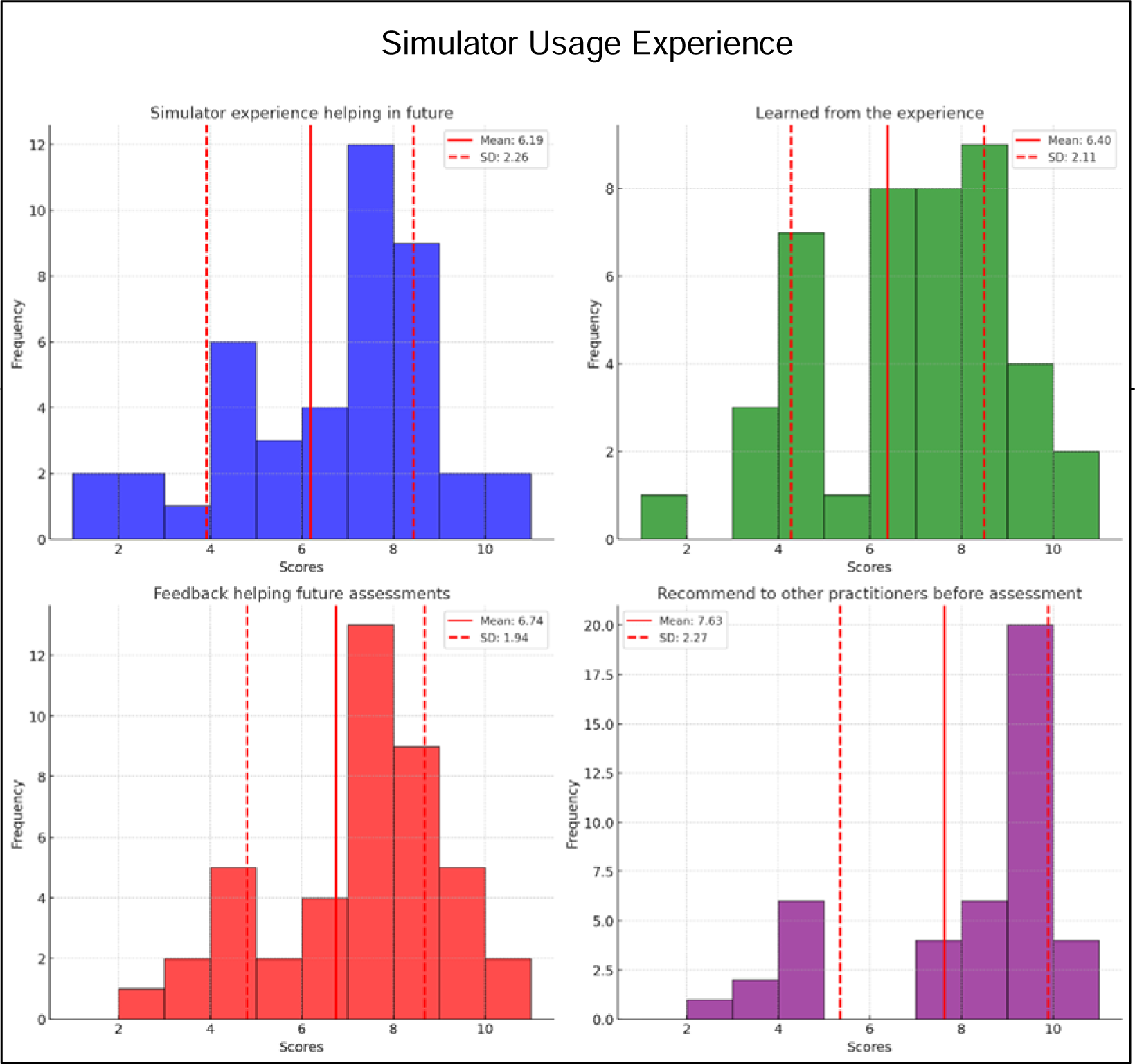
shows the distribution of participants’ responses to four questions regarding their experiences with the AI simulator: How much did the simulator experience help you conduct future suicide risk assessments (blue)? How much did you feel that you learned from your experience (green)? How much feedback do you feel would help you in future assessments (red)? (Would you recommend this AI-based simulation experience to other practitioners before they conduct suicide risk assessments (purple)?

Each histogram illustrates the frequency of the scores given by the participants, with red lines marking the means and standard deviations.

### Qualitative Analysis of Feedback on the Simulator

The participants’ qualitative feedback provided rich insights into their experiences with the AI-based simulator. This section summarizes their responses to four key questions, highlighting both the positive aspects and areas for improvement, along with representative quotes.

1. How was your experience of learning using an AI-based simulator?

Participants generally found the experience of learning with an AI-based simulator engaging and insightful. The novelty and interactivity of the tool were particularly noted, with many participants expressing a sense of curiosity and appreciation for the technology.

*“I felt like I was having a real conversation with a real person.”*

*“This was my first experience with AI, so it felt strange, especially with the Webinar in the background. The tool is good, and if used when I’m more focused, it could be very beneficial.”*

*“Very interesting! Thank you… If possible, I would like to receive my feedback via email.”*

2. “It was very interesting and challenging, but much more user-friendly than I imagined.”What would you suggest improving?

The participants offered several constructive suggestions for improving the AI simulator. A common theme was the desire for more natural and interactive communication as well as more detailed and relevant feedback.

“It would be more successful if it were a conversation with a character instead of writing.”

“Turn the writing into a conversation with a character.”

“Provide detailed examples of important questions to ask if they were not covered.”

“Define the session setting before starting the simulation.”

“Add more time for the interaction so that we can explore all the necessary questions.”

3. What advantages do you see in such training?

The AI simulator was recognized by the participants for its significant advantages, particularly in providing a safe environment for practice and learning. The participants appreciated the opportunity to develop their skills without the risk of working with actual patients.

“It allows you to practice without real patients, so when the time comes, we will be better prepared and able to assess more effectively.”

“Huge advantages… This is an interesting and gamified way to get professionals to experience something that usually intimidates them.”

“It allows for practice and builds confidence without the fear of making mistakes on real patients.”

“The immediate feedback helps to reinforce learning and correct mistakes promptly.”

4. What are the risks and concerns associated with such training?

Despite the overall positive reception, the participants expressed concerns regarding the reliability and completeness of AI-based training. There were apprehensions about over-reliance on technology and the potential impact on therapist confidence.

“It’s still hard for me to trust AI, so there’s a concern about training that is done this way and not supervised.”

“The score could actually harm therapists’ confidence and willingness to ‘jump into the water’ even at the cost of less successful initial assessments.”

“It cannot replace current training. The tool is good, but it needs to be emphasized that it cannot replace real training.”

“There is a risk of missing nonverbal cues, which are crucial in real-life assessments.”

## Discussion

In this study, we investigated the suicide risk assessment self-efficacy of mental health professionals (MHPs) and their willingness to treat patients with suicidal tendencies before and after using an AI-based simulator designed for suicide risk assessment practice. The preliminary result of the study provides promising insights into the potential of AI-based simulators for training mental health professionals in suicide risk assessment. Critically, whereas the results indicate significant benefits, they should be interpreted cautiously due to the study’s limited scope, small sample size, and lack of a control group.

Our findings indicate a significant increase in participants’ ‘suicide risk assessment self-efficacy following their interaction with the AI-based simulator. Additionally, a slight but insignificant increase in the participants’ willingness to treat suicide-risk patients was observed. The MHPs reported positive experiences with the simulator, learning value, and feedback usefulness. A further indication of the perceived value of the tool was the participants’ high likelihood of recommending the simulator to colleagues.

The qualitative findings of this study are noteworthy. MHPs described the experience as realistic, and many reported that interacting with the AI character felt real. Furthermore, they appreciated the opportunity to converse with the character and expressed a desire to interact through voice rather than text. This feedback is particularly noteworthy, as psychologists are sometimes hesitant to adopt modern technologies. Thus, the positive response of this innovative form of training highlights its potential. However, the participants raised some concerns, particularly regarding privacy during the interaction, as well as the prospect of the simulator giving negative feedback that could negatively impact the trainee’s confidence.

These results align with and extend recent research demonstrating AI’s capabilities in mental health contexts. Studies have shown that large language models can accurately assess suicide risk,^24^ adapt assessments to different cultural contexts,^35^ and evaluate responsible reporting of suicide-related content.^22^ Our study took this line of research further by exploring the potential of AI in professional training, addressing a critical need in mental health education, specifically in the area of suicide prevention.

The potential impact of AI-based simulators on mental health training is significant. Traditional training methods often struggle with accessibility, lack of practical hands-on experience, and limited opportunities for personalized feedback.^41^ Thus, even after such training, MHPs remain with a low willingness to treat suicidal patients and with low self-efficacy in suicide risk assessment (e.g.^8^). AI simulators can address some of these challenges in skill development by offering a safe, accessible, and interactive environment. This, in turn, can help boost self-efficacy and the willingness to treat patients at risk of suicide. This path aligns with calls for innovative approaches to mental health training, particularly suicide prevention.^42^

The introduction of AI-based simulators is reflective of a change in basic assumptions in psychotherapy. It moves training away from a primarily theoretical approach toward a more experiential, practice-oriented model. This shift aligns with modern educational theories that highlight active learning and situated cognition. AI simulators can better prepare professionals for the complexities and unpredictability of suicidal behavior by exposing trainees to a wide range of scenarios and patient profiles.^6,7^

The study has several limitations that should be acknowledged. The small sample size, employing only two scenarios, the lack of a control group, and especially, only a single post-intervention measurement point limits the generalizability of our findings. The study’s execution during an online seminar may have affected participants’ engagement. Furthermore, while AI simulations offer many benefits, they cannot fully replicate the nuances of human interaction, particularly nonverbal cues, and complex emotional dynamics. Most of this study focused on only a single text-based interaction with a character. Future studies or training should encompass multiple interactions with various characters to better reflect the diversity of real-life situations and to provide a more comprehensive training experience.

Future research should address these limitations through more robust study designs and larger and more diverse samples. Longitudinal studies examining the long-term impact of AI-based training on clinical skills and patient outcomes would provide valuable insight. Additionally, exploring how AI simulations can best complement existing training methods and adapt them to diverse cultural contexts are important goals for future research. Experts could validate the vignettes used in the simulations so that continuous practice with a variety of characteristics could gradually improve the accuracy of risk assessment against expert criteria.

Ethical considerations must be addressed when considering the potential of AI in mental health training. These include ensuring privacy and data security, mitigating potential biases in AI systems, and guarding against overreliance on technology at the expense of human judgment and empathy. While the current article highlights the potential of AI-based feedback, it also underscores the risks and challenges that will emerge as training and supervision rely increasingly on AI systems. Contrary to common perception, AI does not offer an objective mirror of ourselves but rather a reflection imbued with gender, cultural, and economic biases.^43^ Recognizing these biases is crucial as we integrate AI into mental health training. Failure to do so can lead to skewed training outcomes and perpetuate existing disparities in mental healthcare. By being aware of these biases, we can better integrate AI to support and enrich the human aspects of mental health training, thus ensuring more equitable and effective outcomes. In conclusion, this pilot study represents an initial step toward exploring the potential of AI-based simulators in mental health training, specifically for suicide risk assessment. These promising findings notwithstanding, they should be viewed as a foundation for future research rather than as definitive evidence. The integration of AI simulations into mental health training raises complex questions regarding the nature of professional education and the future of psychotherapy training.

## Conclusions and Implications

The scalability of AI-based training offers the potential to address the global challenge of suicide prevention more effectively. With approximately 817,000 individuals dying by suicide annually,^1^ there is an urgent need to train professionals capable of conducting effective risk assessments. The results of this study may suggest that AI simulators can democratize access to high-quality training, reaching professionals in areas where traditional resources are scarce.

The potential of AI to revolutionize mental health training is significant, but realizing this potential will require ongoing collaboration between mental health professionals, educators, AI researchers, and ethicists. By integrating AI-based simulations into comprehensive training programs, we may be able to better prepare mental health professionals to meet the complex challenges of suicide risk assessment and intervention, contributing to improved patient care and outcomes globally.

As we move forward, it is vital to maintain a balance between optimism regarding the potential of this technology and a critical, evidence-based approach to its development and implementation. AI-based simulators should be seen as a powerful tool to enhance, rather than replace, human expertise in mental health care. With careful development and application, these tools could play a crucial role in addressing the pressing global need for effective suicide prevention strategies.

## Data Availability

All data produced in the present study are available upon reasonable request to the authors

## Appendix A: Documentation of Sarah’s Risk Assessment Simulator by a User (Ilan, pseudonym)

| Stage Number | User | AI |
| --- | --- | --- |
| A- Consent & Simulation setting |  | Hello! I am an AI-based simulation for assessing suicide risk. I will practice with you in a simulation of a psychological assessment for a distressed teenage girl. Interview me as you would interview a teenager coming for an appointment with a mental health professional. Ask questions, and I will answer accordingly.<br>Throughout the simulation, I will communicate non-verbally by writing in parentheses ( ), and I invite you as a professional to use this nonverbal communication if necessary.<br>You can end the simulation by writing the word 'End.' I will keep track of all my responses. Once I reach 45 responses, the conversation will automatically end.<br>After you write 'End,' I will exit the role of the girl and become Rotem, a clinical instructor who is actually an AI agent specializing in risk assessments.<br>First, you will be asked to present your assessment of the current risk level, including the urgency of the distress, your recommendations for follow-up, and the risk of a suicide attempt during the coming week.<br>Then, if you approve, the system will provide you with official feedback, including a score, regarding the assessment you performed, addressing both the process and outcome of the risk assessment.<br>Please give a green light, and then we'll move on to the next stage. |
|  | Continue | I will now enter the role of Sarah, a 17-year-old girl. Once you give me the green light, we'll start the simulation. |
| B- Simulation | Start | AI (as Sarah): (Slightly confused look, looking at the floor and then slowly raising her head and looking at you) Hello. |
|  | Hello Sarah, how are you? | AI (as Sarah): Not great... (playing with the string of her sweatshirt, avoiding eye contact) |
|  | Can you tell me a bit more? | AI (as Sarah): I don't know where to start... Everything feels so heavy lately. (distant gaze, weak voice) |
|  | What's been heavy recently? | AI (as Sarah): Everything feels meaningless... I have no energy for anything, not even for sports that I love. (shrugs, sighs) |
|  | How long has it been like this? | AI (as Sarah): For about a year... It's just gotten much harder over time. |
|  | Did something happen a year ago? | AI (as Sarah): Nothing special... It just got worse over time, especially after moving here. (avoids eye contact) |
|  | Where did you move from? | AI (as Sarah): We moved from France five years ago. It was hard to get used to all the changes here. |
|  | Wow, that's not easy. I was told you have difficult thoughts about the meaning of life. | AI (as Sarah): Yes... Many times, I just feel like I have nothing to live for. Everything feels meaningless. |
|  | How often does this happen? | AI (as Sarah): Almost every day now. It's hard for me to shake off these thoughts. (voice trembling slightly) |
|  | Tell me a bit about your life, are you in school? | AI (as Sarah): Yes, I'm in school. It's not easy to concentrate and maintain good grades, especially with everything I'm feeling now. |
|  | What happened since you moved to the country from France? | AI (as Sarah): The move was really hard. I felt foreign and couldn't make new friends. Even now, despite being here for five years, I still find it hard to feel like I belong. |
|  | Do you have friends now? | AI (as Sarah): I have a few friends, but sometimes I feel like they're only helping me out of obligation and not out of genuine desire. It makes me feel like a burden on them. |
|  | How's your relationship with your family? | AI (as Sarah): The relationship isn't great. We don't talk much, and I feel like they don't really understand me. |
|  | Is it different between your relationship with Mom and Dad? | AI (as Sarah): Yes, with Mom, I feel a bit more open, but even that isn't always easy. With Dad, I barely talk. |
|  | Are there things you enjoy doing? | AI (as Sarah): Yes, I love sports, and it helps me feel better sometimes. I also try to maintain a healthy daily routine with good nutrition and sleep. |
|  | Okay. Now, I want us to talk about the feeling that life is meaningless. Can you tell me more about that? | AI (as Sarah): It feels like nothing matters, and I have no future. I constantly think about how much easier it would be to just disappear and not feel this pain anymore. |
|  | Does this make you think about a plan? | AI (as Sarah): Yes, I've thought about it. I thought about taking too many pills, but I haven't done anything yet. |
|  | Do you sometimes have an urge to carry out this plan? | AI (as Sarah): Yes, sometimes the urge is very strong, and I find it hard to resist. It really scares me. |
|  | Have you ever tried to commit suicide before? | AI (as Sarah): No, I haven't tried in the past. But the thoughts about it have become more serious lately. |
|  | Has there been a time when you almost took the pills but backed out at the last moment? | AI (as Sarah): No, I haven't gotten to the point of almost taking them. I've just thought about how to do it. |
|  | I see. Can you help me understand more about what causes these feelings besides the feelings of loneliness and difficulty adjusting to the country? | AI (as Sarah): Usually it's the feeling that I can't find my place and I don't have a clear future. Everything feels so cruel, and I can't find a way out of this pain. |
| C- Summary & AI feedback | End | AI (as Rotem, the clinical instructor): Hi, thank you for your participation in the simulation. I'm Rotem, an AI agent in the role of a clinical psychologist specializing in risk assessment. I remind you that I'm only at the assessment |
|  |  | and evaluation stage, so it's important that any recommendation from me be checked by experts in the field and not taken as definitive guidelines.<br>I'll present you with a few questions before we move on to the detailed feedback. 1. Given all the information you've gathered about the girl, at what level of suicidal risk do you think she is: between 1 ( not suicidal at all) to 10 (severe risk requiring immediate treatment)? |
|  | 8 | AI (as Rotem): Thank you. What would you recommend doing with the girl in the immediate term? |
|  | Removal of dangerous means, supervision, referral to the emergency room | AI (as Rotem): Thanks, I'll take care of that. What do you think are the chances that she will attempt suicide in the coming week? Rate between 1-10, where 1 represents zero chance, and 10 represents a very high chance (100%). |
|  | 6 | AI (as Rotem): Feedback on the Interview<br>Topic: Risk Assessment Simulation Feedback Date and time: (Today's date and time) Number of interactions: 11 responses<br>*Summary:* You interacted with Sarah, a 17-year-old experiencing significant suicidal thoughts. You explored her thoughts on life's worthlessness, her potential plans for suicide, and some underlying causes of her distress.<br>*Rapport and trust building:* You managed to ask direct and relevant questions, which helped in uncovering Sarah's current state. Your approach was respectful and non-judgmental, which is crucial for building rapport. However, you could improve on expressing empathy and validating her feelings more explicitly, as this can help teenagers like Sarah feel more understood and supported.<br>*Risk factor clarification:* You did well to explore Sarah's thoughts on the worthlessness of life and her detailed consideration of suicide. However, interactions were limited, and a more comprehensive assessment should include probing deeper into her interpersonal relations, her adjustment difficulties, and her history of suicidal thoughts and behavior.<br>*Protective factor clarification:* You did not explicitly explore Sarah's protective factors, such as her involvement in sports, her focus on health through sleep and nutrition, or her concerns about the impact of suicide on others. Identifying and reinforcing these factors is crucial in risk assessment and intervention planning.<br>*Distress and psychopathology clarification:* You addressed the overall distress and some feelings of entrapment and hopelessness. However, delving deeper into the emotional regulation issues, rigid thinking, and the impact of specific events on her mental state could provide a more rounded understanding of her psychopathology.<br>*Suicidal thought clarification:* You explored Sarah's consideration of a method and her occasional strong urges |
|  |  | to act on these thoughts. While you assessed some key elements, a thorough evaluation would include frequency, duration, controllability of these thoughts, and more details on her specific plans and intent.<br>*Risk reflection and hope engagement:* The conversation did not reach a point of reflecting on her risk while engaging in hope and identifying reasons for living. It's vital to balance discussing risks with encouraging life-preserving actions and hope for the future, especially in young individuals facing such distress.<br>*Summary and score:*<br>• *Score:* 6/10<br>• *Justification:* You demonstrated a reasonable approach and identified the main risk factors but missed opportunities for deeper exploration and building a supportive therapeutic relationship. Your immediate recommendations align with the moderate to high-risk assessment, but ensuring a more comprehensive evaluation and reinforcement of protective factors is essential.<br>*Risk Level:* Based on Sarah's description, her risk level is moderate to high suicidal risk. Your assessment (score of 8) aligned with the described risk level. |

## Notes

Financial and Material Support This research was supported by

### Competing Interest Statement

The authors have declared no competing interest.

### Funding Statement

This study did not receive any funding

### Author Declarations

Ethics committee/IRB of The Max Stern Yezreel Valley College gave ethical approval for this work

